# Early Morning Checkpoint Inhibitor Infusion and Overall Survival of Patients with Metastatic Cancer: an In-depth Chronotherapeutic Study

**DOI:** 10.1101/2023.09.06.23295010

**Authors:** Simona Catozzi, Souad Assaad, Lidia Delrieu, Bertrand Favier, Elise Dumas, Anne-Sophie Hamy, Aurélien Latouche, Hugo Crochet, Jean-Yves Blay, Jimmy Mullaert, Annabelle Ballesta, Pierre Heudel

## Abstract

**Introduction:** Recent retrospective studies suggest potential large patient’s benefit through proper timing of immune checkpoint inhibitors (ICI). The association between ICI treatment timing and patient survival, neoplastic response and toxicities was investigated, together with interactions with performance status (PS) and sex.

**Methods:** A cohort of patients with metastatic or locally advanced solid tumors, who received pembrolizumab, nivolumab, atezolizumab, durvalumab, or avelumab, alone or with concomitant chemotherapy, between November 2015 and March 2021, at the Centre Leon Bérard (France), was retrospectively studied.

**Results:** 361 patients were investigated (80% non-small cell lung cancer patients, mean [SD] age: 63 [11] years, 39% of women, 83% PS0-1 at first infusion, 19% received concomitant chemotherapy). ICI were administered from 07:25 to 17:21 and optimal morning/afternoon cut-off was 11:37. Morning infusions were associated with increased OS as compared to afternoon (median 30.3 vs 15.9 months, p=0.0024; HR 1.56 [1.17-2.1], p=0.003). A strong PS-timing interaction was found (PS0-1 patients, HR=1.53 [1.10-2.12], p=0.011; PS2-3 patients, HR=0.50 [0.25-0.97], p=0.042). Morning PS0-1 patients displayed increased OS (median 36.7 vs 21.3 months, p=0.023), partial/complete response rate (58% vs 41%, p=0.027), and grade1-3 toxicities (49% vs 34%, p=0.028). Mortality risk ratio between infusions at worst time-of-day, estimated at 13:36 [12:48-14:23], and in early morning was 5.5-fold ([2-20], p=0.008). Timing differences in toxicities resulted significant only in female patients (women vs men: p<0.001 vs 0.4).

**Conclusions:** Early morning ICI infusion was associated with increased OS, response, and toxicities in patients with PS0-1 as compared to later infusions within the day. Prospective randomized trials are needed to confirm this retrospective study.

## Introduction

Chronotherapy is based on the principle that a drug can have different activity depending on its infusion schedule over 24h as a result of circadian rhythms in most physiological functions of the organism^1^. Large timing-related differences in drug antitumor efficacy and/or toxicities have been demonstrated for more than 50 anticancer compounds in mouse studies^2,3^. A recent review analyzed 18 randomized clinical trials in cancer patients, 11 of them reporting a significant decrease in toxicities in the arm with optimized chemotherapy administration timing, with a maintained antitumor efficacy^4^. Importantly, three prospective clinical trials reported an increase in survival outcomes and/or response rates while none of these 18 studies showed a decrease in efficacy. Cancer chrono-chemotherapy is most documented for colorectal cancer patients receiving 5-fluorouracil-leucovorin, in combination with oxaliplatin and/or irinotecan for which proper sex-specific drug timing reduced the incidence of toxicities by up to five-fold, while leading to maintained or improved neoplastic control and/or patient survival^3,5–11^.

Over the past decade, immune checkpoint inhibitors (ICI) have completely changed the paradigm for the treatment of locally advanced or metastatic cancers and dramatically improved patient overall survival^12,13^. Administered as a monotherapy or in combination with chemotherapy, ICI improve tumor response, yet at the cost of adverse events which, although infrequent, are sometimes irreversible or even fatal^14–16^. Recently, eight retrospective studies suggested that the time-of-day of ICI infusion may impact the survival of patients with metastatic cancer ^17–24^. In these different populations, including patients with metastatic lung cancer, advanced melanoma, renal cell or urothelial cancers, progression-free and/or overall survival (OS) were longer when most of the immunotherapy infusions were performed in the early part of the day, with median OS being lengthened by up to 3.6-fold in the “morning” vs “afternoon” patient groups.

To consolidate these results, we retrospectively evaluated the impact of ICI administration timing on patient survival, neoplastic response, and toxicities in a large population of patients with advanced cancers. We investigated possible interactions with the patient performance status (PS), a major predictor of patient OS which has been positively associated with circadian disruption^1,26^, together with sexual dimorphism as previously reported for cancer chronotherapies^1,25^. From a methodological point of view, we propose a rationale for optimizing morning/afternoon group cut-off and further present a periodic mortality risk model to precisely predict optimal infusion time-of-day.

## Materials and methods

### Study design and cohort population

This retrospective study was approved in July 2020, by the local data protection officer, on behalf of French regulatory authorities in accordance with MR004 methodology (R201-004-207). All patients were informed of the possibility of their health data being used for research purposes and expressed no opposition to this possibility. The patient cohort was selected with ConSoRe, a data mining solution^27^. This tool was used to find all patients over the age of 18 treated with at least one course of immunotherapy at the Centre Leon Berard (Lyon, France) between November 2015 and March 2021 for unresectable locally advanced or metastatic lung cancer and between December 2016 and February 2020 for other types of metastatic solid cancers. We excluded patients treated with (i) immunotherapies other than pembrolizumab, nivolumab, atezolizumab, durvalumab, avelumab, (ii) ipilimumab in monotherapy, (iii) combinations of ipilimumab with nivolumab or durvalumab with tremelimumab, or (iv) receiving adjuvant or neoadjuvant immunotherapy.

### Patient data collection

Clinical data and tumor characteristics were extracted from electronic medical records including age, sex, body mass index (BMI) at diagnosis, PS (from 0 to 3) at first ICI administration, tumor histology and PDL-1 status (considered positive if >1%), and the number of previous systemic treatments.

### ICI treatment and timing

Nivolumab (240 mg q2week), pembrolizumab (200 mg q3week), atezolizumab (840 mg q2week or 1200 mg q3week), durvalumab 1(0 mg/kg of body weight q2week or 1500 mg q4week), and ipilimumab (1 or 3 mg/kg q3weeks) were administered intravenously. ICI timing slots were randomly allocated for each course by the day-hospital coordinator. For each immunotherapy infusion, the clock time of arrival of the immunotherapy solution in the day hospital from the pharmacy was automatically annotated by computer. An internal audit performed annually confirmed that the actual drug administration to the patient systematically occurred within the next 20 minutes after drug solution arrival on average, with a maximum delay of one hour.

### Outcomes

The primary outcome was OS, determined as the time between the date of first ICI infusion and the date of death of any cause or date of last news. Secondary outcomes were the rate of complete response and occurrence of toxicities. Evaluation of stable disease, partial and complete response rate was performed by RECIST 1.1. Toxicities were based on adverse event occurrence according to criteria NCI CTCAE v5.0. We excluded toxicities not related to immune adverse.

### Exposure: optimizing timing cut-offs

Firstly, the patient population was dichotomized into “morning” and “afternoon” groups, based on their median infusion times being, respectively, before and after the specified timing cut-off (i.e., patients having received >50% of their ICI infusions before/after the cut-off). The predictiveness curve method was used to estimate the optimal timing cut-off that better separates morning/afternoon groups in terms of OS. It is based on a risk model specified as a univariable Cox proportional hazard regression^28^.

For dividing the cohort into four patient groups regarding timing, we computed the restricted mean survival times (RMST, i.e. areas under the survival curves) up to time t=1250 days, which corresponded to the time when ∼90% of events had occurred^29^. Optimal group cut-offs which maximized the pairwise RMST differences between groups were obtained by grid search, provided a minimal group size of 30 patients (i.e., 10% of the PS0-1 population).

### Statistical Analysis

For continuous variables, we reported mean/SD or median/interquartile range, following whether they fulfilled the normality assumption or not, respectively. For categorical variables, absolute values and proportions are provided. Statistical differences between patient groups were assessed with T-test/Kruskal-Wallis for normally/non-normally distributed continuous variables respectively, Pearson’s chi2 test or Fisher’s exact test for categorical variables, and Wilcoxon rank test for ordered categorical variables (here tumor response, number of toxicities and highest toxicity grade). Association of categorical variables (here, sex and timing) with ordered categorical variables (response and toxicities) was tested using ordinal regression (*clm* function in ordinal R package). Missing values were handled with the MICE algorithm, using the classification and regression tree (*cart*) method for imputation (*mice* R package).

The OS was estimated by Kaplan-Meier curves. Median follow-up was calculated with the reverse Kaplan-Meier estimator. Differences between patient group survival were evaluated with a global log-rank test or a Peto log-rank test for under-populated and unbalanced PS2-3 patient subgroups^30^. Univariable and multivariable Cox models were used to estimate the impact of patient and treatment characteristics on survival.

For PS0-1 patients, a sinusoidal Cox regression model was used to investigate the association of ICI timing on treatment outcomes, building on the continuous cyclic nature of the timing variable^31^. The 24h-periodic risk model is characterized by two parameters: an acrophase (time of peak) and an amplitude (half-distance between minimum and maximum, see Supplementary Information). The variance and significance of these parameters were calculated using the delta method^31^. The goodness-of-fit of the periodic model was evaluated by the calibration curve method (*calPlot* function in R)^32^. The significance level for all tests was set at 0.05. All computations were done using R v4.2.2 and Python 3.10.4.

## Results

### Description of the patient population and ICI treatments

Out of the 385 patients eligible for this study, 24 patients for whom more than 20% of ICI infusion timing was not available were excluded from the cohort. Thus, this study comprised 361 patients with metastatic cancer, including 289 (80%) non-small cell lung cancer patients (mean age 62.5 years, mean BMI 23.8, men proportion 61.5%, Table 1). The number of patients with PS of 0, 1, 2, and 3 were, 67 (18.6%), 231 (64.2%), 53 (14.7%), and 9 (2.5%) patients, respectively. Prior to ICI, 16 (4%) patients received immunosuppressive therapy for an autoimmune disease, mostly sarcoidosis, Crohn’s disease/ulcerative colitis and lupus erythematosus.

**Table 1.** Patient and tumor characteristics. The time cut-off dividing morning and afternoon patient groups was set to 11:37.

|  |  | Missing | Overall | Morning | Afternoon | P-Value |
| --- | --- | --- | --- | --- | --- | --- |
| n |  |  | 361 | 136 | 225 |  |
| Median infusion time, median [Q1,Q3] |  | 0 | 12.3 [10.9,15.3] | 10.6 [9.8,11.1] | 14.9 [12.7,15.7] | <0.001 |
| Age (years), mean (SD) |  | 0 | 62.5 (10.7) | 61.7 (11.0) | 62.9 (10.5) | 0.324 |
| BMI, mean (SD) |  | 2 | 23.8 (4.5) | 24.2 (4.6) | 23.6 (4.4) | 0.210 |
| Sex, n (%) | Female | 0 | 139 (38.5) | 48 (35.3) | 91 (40.4) | 0.388 |
|  | Male |  | 222 (61.5) | 88 (64.7) | 134 (59.6) |  |
| Performance status, n (%) | 0-1 | 1 | 298 (82.8) | 124 (91.9) | 174 (77.3) | 0.001 |
|  | 2-3 |  | 62 (17.2) | 11 (8.1) | 51 (22.7) |  |
| Immunoallergic history, n (%) | No | 1 | 337 (93.6) | 122 (90.4) | 215 (95.6) | 0.085 |
|  | Yes |  | 23 (6.4) | 13 (9.6) | 10 (4.4) |  |
| Histological type, n (%) | NSCLC | 0 | 289 (80.1) | 113 (83.1) | 176 (78.2) | 0.060 |
|  | Colorectal |  | 3 (0.8) | 1 (0.7) | 2 (0.9) |  |
|  | Melanoma |  | 18 (5.0) | 4 (2.9) | 14 (6.2) |  |
|  | ENT |  | 22 (6.1) | 3 (2.2) | 19 (8.4) |  |
|  | Breast |  | 1 (0.3) | 1 (0.7) |  |  |
|  | Urinary |  | 27 (7.5) | 14 (10.3) | 13 (5.8) |  |
|  | Pancreas |  | 1 (0.3) |  | 1 (0.4) |  |
| Tumor PDL1 expression, n (%) | >1 | 168 | 68 (35.2) | 27 (38.0) | 41 (33.6) | 0.612 |
|  | <1 |  | 51 (26.4) | 20 (28.2) | 31 (25.4) |  |
|  | Not assessed |  | 74 (38.3) | 24 (33.8) | 50 (41.0) |  |
| Previous systemic treatment, n (%) | No | 1 | 126 (35.0) | 51 (37.8) | 75 (33.3) | 0.458 |
|  | Yes |  | 234 (65.0) | 84 (62.2) | 150 (66.7) |  |
| Type of first immunotherapy, n (%) | Atezolizumab | 0 | 20 (5.5) | 10 (7.4) | 10 (4.4) | 0.189 |
|  | Durvalumab |  | 22 (6.1) | 11 (8.1) | 11 (4.9) |  |
|  | Nivolumab |  | 132 (36.6) | 41 (30.1) | 91 (40.4) |  |
|  | Pembrolizumab |  | 186 (51.5) | 74 (54.4) | 112 (49.8) |  |
|  | Ipilimumab |  | 1 (0.3) |  | 1 (0.4) |  |
| Concurrent chemotherapy, n (%) | No | 0 | 294 (81.4) | 107 (78.7) | 187 (83.1) | 0.363 |
|  | Yes |  | 67 (18.6) | 29 (21.3) | 38 (16.9) |  |
| Two sequential ICI, n (%) | No | 0 | 351 (97.2) | 132 (97.1) | 219 (97.3) | 1.000 |
|  | Yes |  | 10 (2.8) | 4 (2.9) | 6 (2.7) |  |
| Line of immunotherapy treatment, n (%) | 1st line | 1 | 127 (35.3) | 51 (37.8) | 76 (33.8) | 0.679 |
|  | 2nd line |  | 188 (52.2) | 65 (48.1) | 123 (54.7) |  |
|  | 3rd line |  | 33 (9.2) | 14 (10.4) | 19 (8.4) |  |
|  | 4th line |  | 11 (3.1) | 5 (3.7) | 6 (2.7) |  |
|  | 5th line |  | 1 (0.3) |  | 1 (0.4) |  |
| Treatment with anti-inflammatory drugs, n (%) | No | 1 | 344 (95.6) | 125 (92.6) | 219 (97.3) | 0.064 |
|  | Yes |  | 16 (4.4) | 10 (7.4) | 6 (2.7) |  |
| Next treatment, n (%) | No | 2 | 155 (43.2) | 58 (43.3) | 97 (43.1) | 1.000 |
|  | Yes |  | 204 (56.8) | 76 (56.7) | 128 (56.9) |  |
| Type of next treatment, n (%) | Chemiotherapy | 157 | 168 (82.4) | 61 (80.3) | 107 (83.6) | 0.691 |
|  | Surgery |  | 1 (0.5) | 1 (1.3) |  |  |
|  | Immunotherapy |  | 4 (2.0) | 2 (2.6) | 2 (1.6) |  |
|  | Radiotherapy |  | 14 (6.9) | 6 (7.9) | 8 (6.2) |  |
|  | Targeted therapy |  | 17 (8.3) | 6 (7.9) | 11 (8.6) |  |

The types of ICI received by the patients were pembrolizumab (51.5% of all infusions), nivolumab (37%), atezolizumab (5.5%), durvalumab (6%), and ipilimumab (0.3%). None of the included patients received avelumab. ICI were administered as a first line of treatment for 127 (35%) cancer patients, as a second line for 188 (52%) and later on for the 44 (13%) remaining patients. Overall, 67 patients were receiving concurrent chemotherapy, and 10 received two sequential different immune checkpoint inhibitors.

ICI were administered from 07:25 to 17:21, with peaks of hospital activity in the late morning and mid-afternoon (Figure S1a). The variability in shipping times per patient varied by only 90 min on average (IQR [48-120] min), attesting for relatively homogeneous ICI administration timing for a given patient (Figure S1b).

### Defining the optimal morning/afternoon cut-off

The *optimal* timing cut-off that better separated the morning and afternoon infusion groups in terms of OS was estimated at 11:37 (see Methods; Figure S2). It provided a better morning/afternoon dichotomization than the median of the patients’ median ICI infusions times here equal to 12:18 (Figure S2; optimal vs median: ΔRMST=160 vs 120 days, log-rank p=0.002 vs 0009). All patient characteristics were similar between morning and afternoon infusion groups defined using the optimal cut-off, apart from the PS, as a larger proportion of PS2-3 patients was shown in the afternoon population (p=0.001; Table 1). Both groups were also balanced for all variables but PS when restricting the analysis to lung cancer patients only (p=0.02, Table S1).

### Treatment duration, Response and Toxicities in morning vs afternoon groups, and interaction with PS

Patients received a median number of 8 courses (range: 1 to 107), for a total number of 4049 infusions, and a median duration of 98 days. Forty-one patients (11%) discontinued their treatment, mostly because of progressive disease (Table 2). Stable disease was observed in 114 (32%) patients, and either partial or complete response in 167 (47%) patients (Table 2). Toxicities were developed by 143 (40%) patients, thereof 45 (32%) underwent at least 2 toxic events and 23 (16%) suffered from grade 3-4 toxicities (Table 2).

**Table 2.**
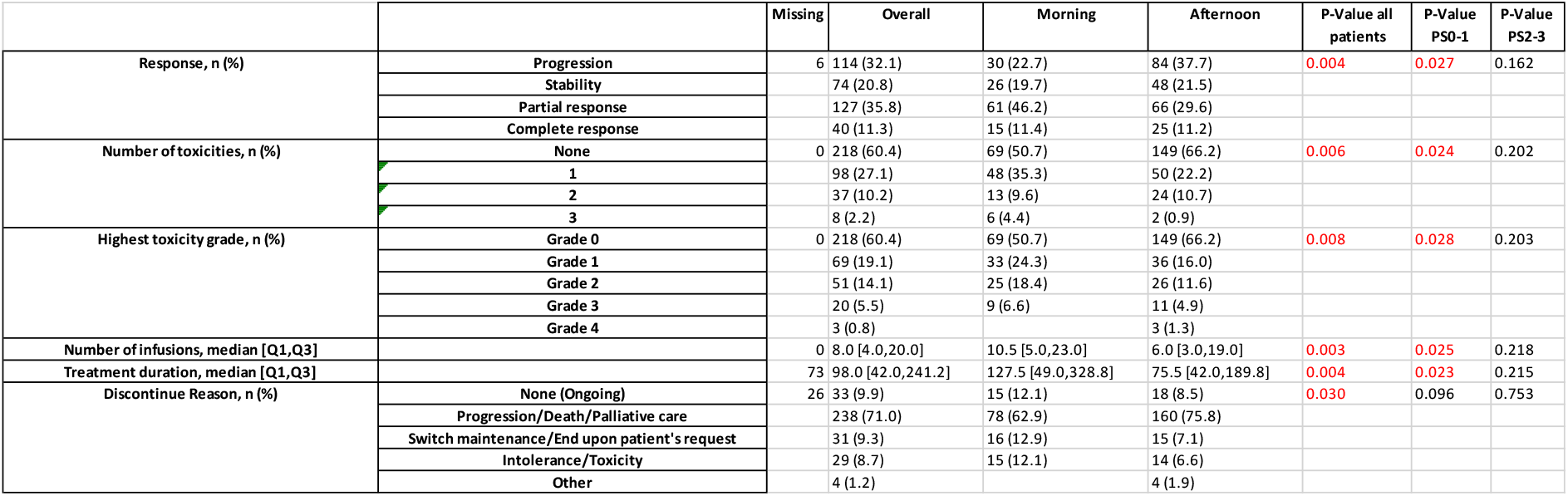
Treatment outcomes. The time cut-off dividing morning and afternoon patient groups was set to 11:37. The reported types of toxicity were cardiac, skin, diabetic, digestive, endocrine, hepatic, hypophysic, nephritic, neurological, ophthalmic, pancreatic, pulmonary, renal, rheumatological, thyroidal; with grade from 1 to 4.

Morning patients received a higher number of infusions over a longer treatment period and responded significantly better to the ICI treatment as compared to the afternoon group, but also developed more toxicities (Table 2). These results still held when analyzing the lung patients only (Table S2). Treatment response and tolerability timing-related differences observed for the whole population were also significant for PS0-1 patients but were systematically not validated for PS2-3 ones (Table 2). A sexual dimorphism was observed for number of toxicity (p=0.036), worst toxicity level (p=0.04687) but not for response and no interactions between sex and timing were validated. Morning superiority over afternoon in terms of ICI efficacy observed for the whole population was even more significant in female patients while being at the limit of significance in men (women vs men: treatment duration p=0.028 vs 0.045; number of infusions p=0.006 vs 0.09, response p=0.032 vs 0.065) (Table S3). Strikingly, ICI timing considerably affected treatment tolerability in women while being not a predictor variable in men (women vs men: number of toxicities p<0.001 vs 0.4, highest toxicity grade p=0.003 vs 0.62).

### Dichotomized morning/afternoon survival analysis revealed superiority of morning ICI infusions in patients with PS0-1, but not with PS2-3

Median follow-up was 32.2 months (95% CI: 12.0–42.6), and median OS was 21.1 months, with 213 deaths occurring over 361 patients. Morning patients showed a median OS almost twice as long as that of the afternoon patients (30.3 versus 15.9 months, p=0.0024; Figure 1a). According to a univariable analysis, the mortality risk was increased by afternoon infusions as compared to morning ones (HR=1.56, 95% CI: 1.17-2.10, p=0.003), PS2-3 as compared to PS0-1 (HR=3.82, 95% CI: 2.79-5.23, p<0.001) and ICI administration as a second or later line (HR=1.46, 95% CI: 1.08-1.98, p<0.013, Figure 1b). The superiority of morning vs afternoon infusions regarding OS was observed for all patient categories, except for the PS2-3 patient subset, yet only significantly for the lung cancer subset, women, patients in second or higher line of treatment, and patients not receiving concomitant chemotherapy which displayed the opposite trend (Figure 1c). For the PS0-1 patient subset, the only univariable predictive factor was the administration timing which remained significant in lung cancer and women subgroups only (Figure S3). For the lung cancer subpopulation only, PS0-1 patient median OS was also longer in the morning group (morning vs afternoon: not reached vs 23.3 months, log-rank p=0.016, Figure S4). For PS2-3 lung cancer patients, median OS was similar between both timing groups (morning vs afternoon: 2.4 vs 3.4 months, Peto log-rank p=0.197).

**Figure 1.**
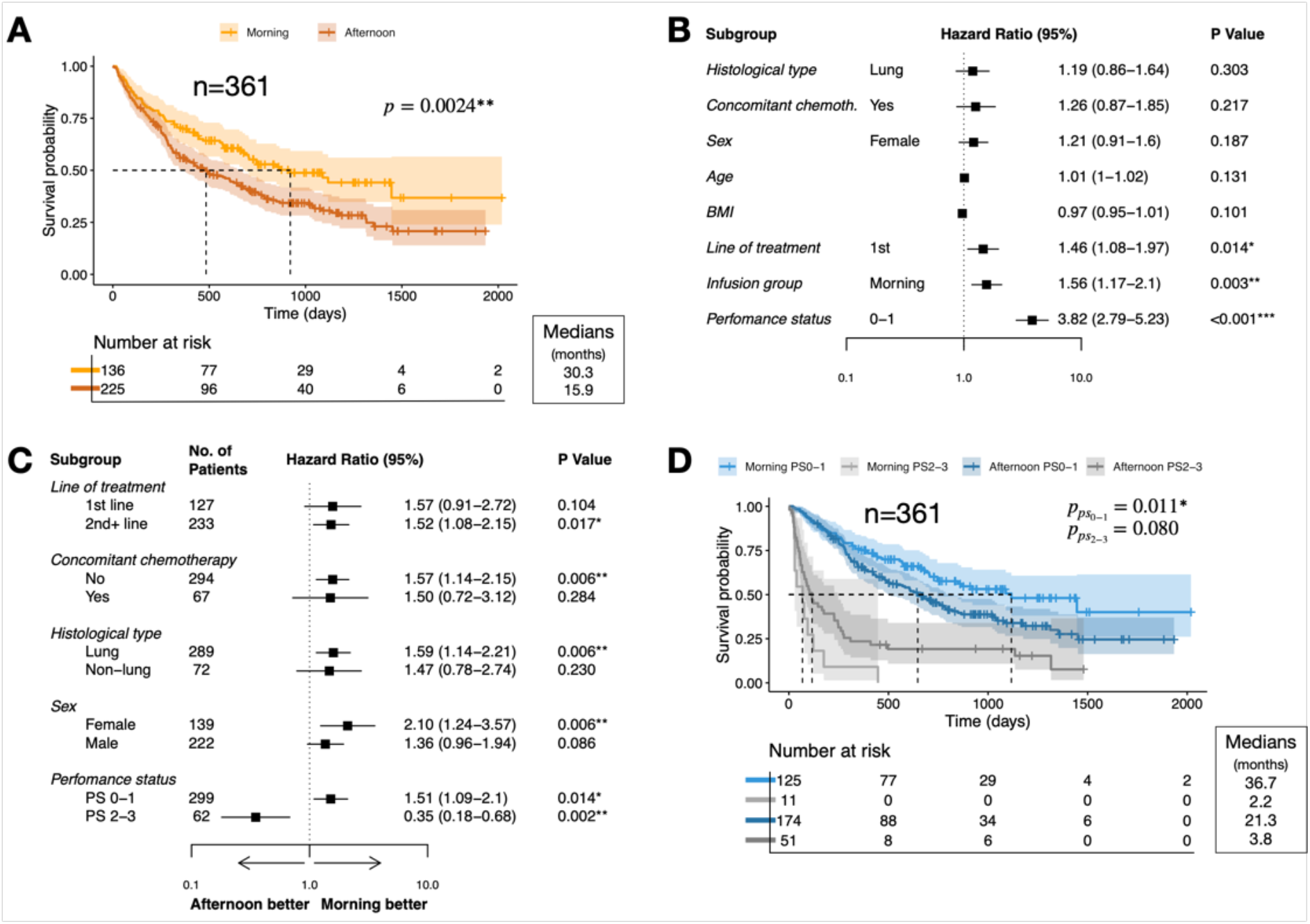
Association between ICI infusion timing and patient OS. (A) OS of morning and afternoon infusion groups and log-rank test (B) Forest plots of OS HRs, according to patient characteristics. HRs and 95% confidence intervals from univariable Cox models are reported. (C) Forest plot of OS HRs by patient subgroup. HRs and 95% confidence intervals of an earlier death for afternoon versus morning infusion groups (univariable Cox models). (D) OS of morning and afternoon infusion groups stratified by PS. Significance was evaluated with log-rank test for PS0-1 and with Peto’s log-rank test for PS2-3.

Since PS was a strong prognosis factor and that the morning/afternoon groups were imbalanced for this variable (Table 1), we investigated PS-stratified survival curves (Figure 1d). Patients with PS0-1 had a significant increase in median OS while receiving ICI in the morning as compared to afternoon (36.7 vs 21.3 months, p=0.011). On the contrary, PS2-3 patient survival was not significantly impacted by administration timing, although the 11 patients of the morning group displayed a shorter OS as compared to the 51 patients who received afternoon infusions (p=0.08). The effect of timing, sex, and line of treatment, was investigated through a multivariable Cox model stratified on PS, that included the pairwise interactions between these three factors as well as their interaction with PS. The only significant variables were timing (p=0.04) and PS-timing interaction (p=0.003). Afternoon infusions increased mortality risk in PS0-1 patients (HR=1.53, 95% CI: 1.10-2.12, p=0.011) and decreased it in PS2-3 patients as compared to morning ICI treatment (HR=0.50, 95% CI: 0.25-0.97, p=0.042).

### Deciphering the Periodic Nature of ICI Timing Impact on Overall Survival

As consistent timing effect was not demonstrated for PS2-3 patients, we focused on PS0-1 patients only (n=299). Four patient groups with different ICI timing were computed (see Methods). The optimal subgroups consisted in patients treated in the *early morning* (08:38-09:53), *morning/midday* (9:53-12:40), *early afternoon* (12:40-14:52), and *late afternoon* (14:52-16:58) (Figure 2a,b). A clear distinction was observed between the group of earliest infusions, associated with the longest survival, and the one of early afternoon, resulting in the worst survival (median OS: 47.5 vs 15.9 months), while the other two groups resulted intermediate (global log-rank p = 0.0005). The *early morning* group was significantly superior to the three other groups in terms of OS (pairwise log rank p<0.005, Figure 2b).

**Figure 2.**
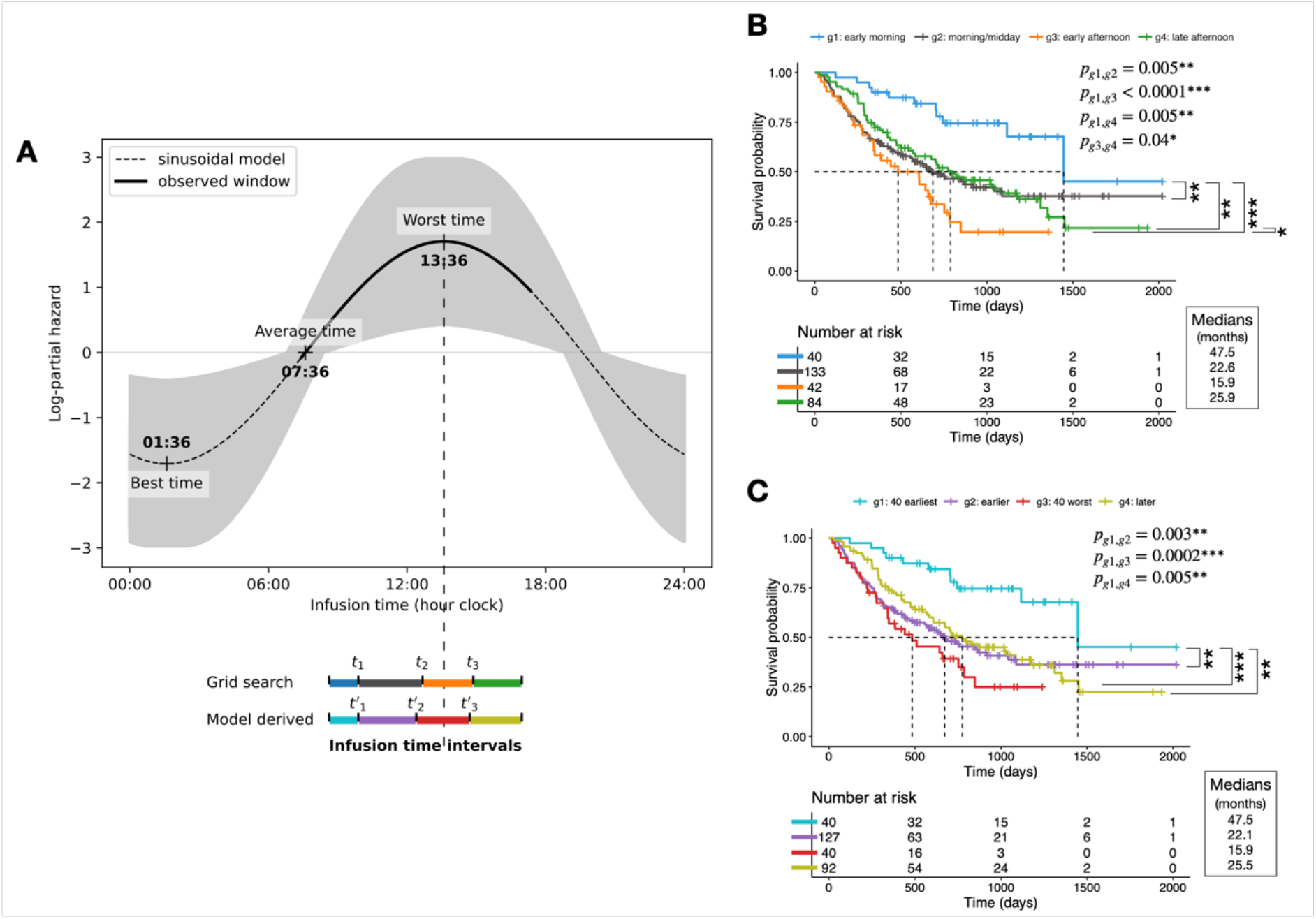
Periodic risk model to analyze OS of patients with PS0-1. (A) Log-partial hazard of the periodic Cox regression model. The shaded area represents the model 95% CI. (B) Kaplan-Meier curves for the four infusion groups for which the difference in RMST is maximized. The timing cut-offs defining the groups are 09:53, 12:40, and 14:52 (denoted as t_1_, t_2_ and t_3_ in panel A). (C) Kaplan-Meier curves for the four infusion groups defined using the periodic model. The timing cut-offs defining the groups are 09:53, 12:24, and 14:43 (denoted as t′_1_, t′_2_ and t′_3_ in panel A).

Such “increasing and then decreasing” pattern in the risk of death for increasing infusion times ruled out linear relationship between both variables and suggested a sinusoidal one, as could have been anticipated from the periodic nature of the infusion timing variable.

Hence, we used a sinusoidal Cox regression model to assess timing impact on the mortality risk (see Methods, Supplementary Information, Figure 2a). This model showed a good agreement with survival data (Figure S5). To quantitatively validate this new model, the PS0-1 patient population was divided into four timing groups, now only using the mortality risk as predicted by the periodic model (Figure 2a and Figure S6). The model-driven survival curves resulted analogous to the ones obtained independently of it (Figure 2b,c), with very similar cut-offs (09:53 vs 9:53, 12:24 vs 12:40, and 14:43 vs 14:52), thus validating the model accuracy.

The validated model predicted *worst* infusion timing to occur at 13:36 (95% CI: 12:48-14:23) with associated mortality risk being 5.5-fold higher than that predicted for infusions at the “average” timing, found 6h earlier, at 07:36 (HR 95% CI: 1.6-19.7, p=0.008). The extrapolated *best* timing would occur 12h earlier, at 01:36 (95% CI 00:48-2:23), although such prediction lacks proper validation due to the absence of survival data for patients treated during the evening/night. The amplitude of timing impact was larger in women as compared to men and not significant in the latter ones (worst vs average timing HR: 2.64 (0.52–4.75) p=0.015 in women, 1.13 (0.44–2.78) p=0.16 in men, Figure S7). However, female and male patients displayed similar worst time: 13:25 (12:36-14:12) vs 13:51 (12:08-15:32), respectively.

Comparatively with the dichotomized morning/afternoon approach, this refined approach could better isolate best and worst ICI timing windows as the difference in median OS between best (early morning or morning) and worst (early afternoon or afternoon) infusion groups were 3 vs 1.7-fold, and the difference in RMST were 444 vs 137 days, respectively (Figure 2b vs Figure 1d).

## Discussion

In a large cohort of patients diagnosed with advanced cancers, we retrospectively investigated the effect of ICI infusion timing on patient survival, neoplastic response, and toxicities. We report for the first time that infusion timing impacted treatment outcomes with a strong interaction with PS. Indeed, morning ICI infusions were consistently associated with longer OS and treatment duration, better response and higher toxicities as compared to later infusions, but only in patients with PS0-1. Grade 1 or 2 toxicities were more often developed among the morning infusion patients, which aligned with their reported association with immunotherapy efficacy^33–35^. For patients with PS2-3, low timing impact might be explained by the dampening or loss of proper circadian organization^1,26^. Our results are in line with the eight recently published studies on cancer chrono-immunotherapies which investigated almost exclusively PS0-1 patients and all concluded on the best timing group being the earliest studied one, either in terms of OS, PFS or response, regardless of the chosen morning/afternoon timing cut-off^17–24^. Here, timing univariable effect was more pronounced in women than in men (Figure 1C, Figure S7). Moreover, large timing-related differences in toxicities were found for women while none for men (Table S3). This finding is in agreement with higher amplitudes in women as compared to men in timing variations of toxicities induced by irinotecan in combination with 5-fluouracil and oxaliplatin for patients with metastatic colorectal cancer^9^.

An essential aspect addressed in this study was the methodology for optimizing timing cut-offs for which no consensus currently exists. In the former chrono-immunotherapy studies, the morning/afternoon groups were chosen either from theoretical considerations or using the median of timing and ranged from 12:55 to 16:30^17–24^. However, the timing cut-off needs to be optimized as it is linked to the optimal treatment timing. To answer this question, we suggested (i) the predictiveness curve methodology to optimally dichotomize the patient cohort, (ii) a more refined approach based on a periodic risk model that provided an estimation of the optimal time-of-day for ICI infusion, and quantified the benefit associated to proper timing in terms of patient OS.

This study confirms a probable large benefit in terms of patient OS from early morning ICI infusions as compared to any other times within day hospital opening hours, thus raising the issue of clinical implementation of such result. This suggests organizing care accordingly by planning patients receiving ICI as early as possible while patients receiving other molecules would be treated later during the day. A clinical trial will be implemented to test this hypothesis.

The periodic model was used to extrapolate the best infusion timing over the 24h window which was predicted during the night, at 01:36. Such finding advocates for technological development (e.g. delayed-release oral drug formulation, programmable delivery pumps) that would allow for ICI infusion outside of hospital opening hours, in order to validate the further increase in patients OS associated with infusions in the second half of the night as compared to early morning infusion. The introduction of remote smart technologies to assist with chronotherapy implementation is desirable to ensure patient’s benefit from chrono-tailored immunotherapy and to free up space in overcrowded day hospitals.

The main limitation of this study is that it is a retrospective investigation based on a monocentric cohort aggregating patients with different cancers or lines of treatment.

Our results confirmed an association between early morning immunotherapy infusions and increased OS in patients with PS0-1, as compared to later infusion timing. Furthermore, new methodologies for studying drug optimal timing were presented. Prospective randomized clinical trials are urgently needed to corroborate our findings and allow for a rapid translation into the clinics.

## Data Availability

The source code generated during this study is available online at https://github.com/semolas/early-morning-checkpoint-inhibitors.

## Acknowledgements

We thank Prof Francis Lévi and Dr Abdoulaye Karaboué for fruitful discussions.

