## Supplementary Figures for "Early Morning Checkpoint Inhibitor Infusion and Overall Survival of Patients with Metastatic Cancer: an In-depth Chronotherapeutic Study"

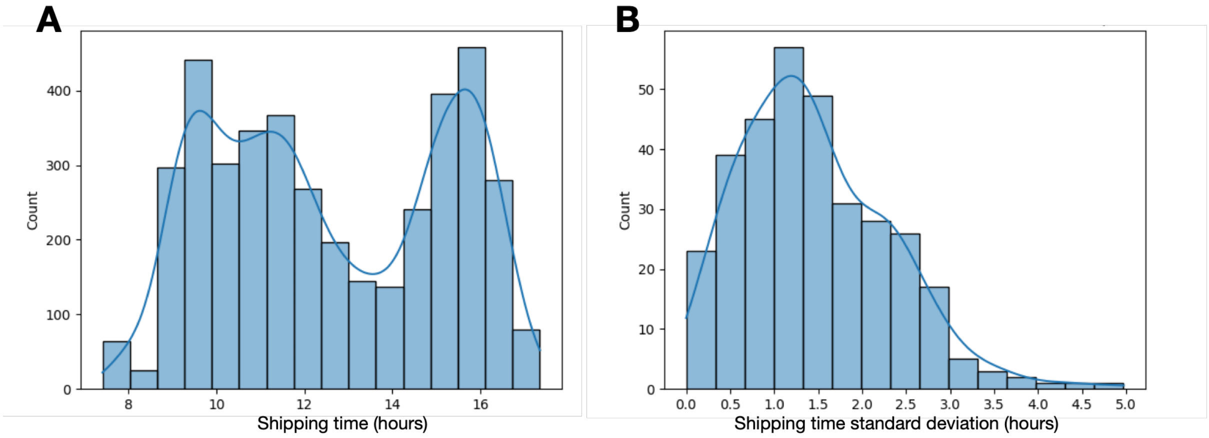


***Figure S1:*** (A) Distribution of the ICI shipping time ranging from 07:25 to 17:21 (4904 data points represented). (B) Distribution of the variance of the ICI shipping time per patient (361 data points). Median (IQR) = 1.3 (0.8-2.0) hours.

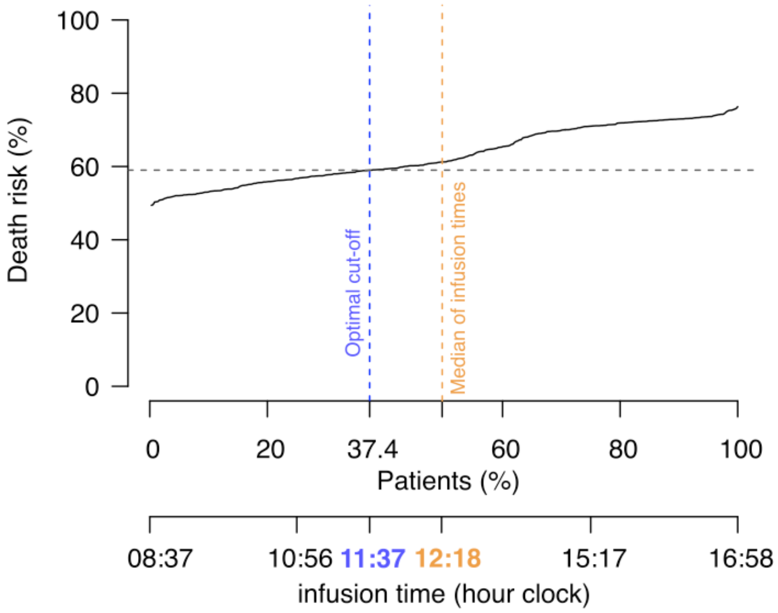


***Figure S2.*** Infusion time cut-off given by the predictiveness curve: optimal cut-off at t=11:37 (blue) and sub-optimal cut-off for the median of the patient median infusion times at t=12:18 (orange).

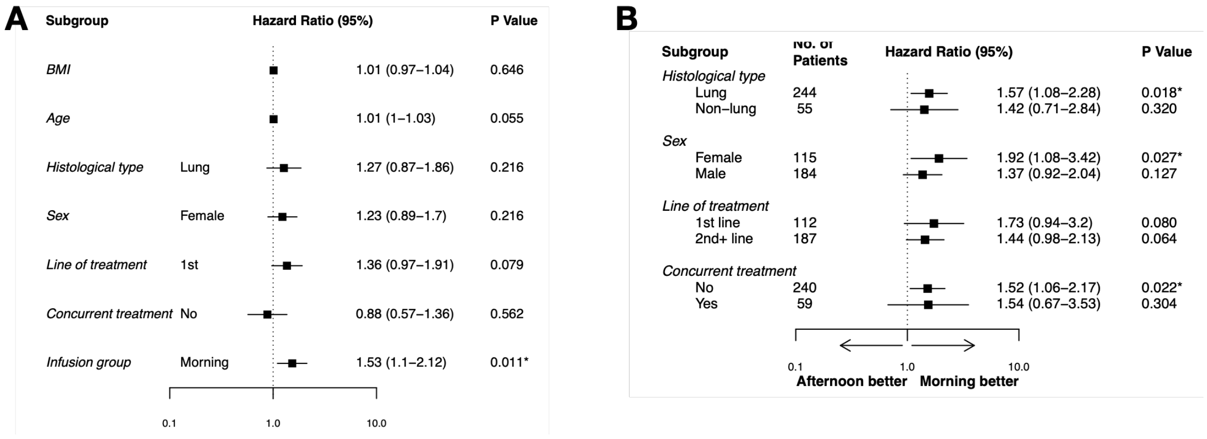


***Figure S3.*** (A) Forest plots of overall survival, according to patient characteristics (univariable Cox models) for the PS0-1 patients only. Hazard ratios and 95% confidence intervals of shorter (HR<1) or longer (HR>1) survival time. (B) Forest plot of overall survival hazard ratios by subgroup for PS0-1 patients only. Hazard ratios and 95% confidence intervals of an earlier death for afternoon versus morning infusion groups.

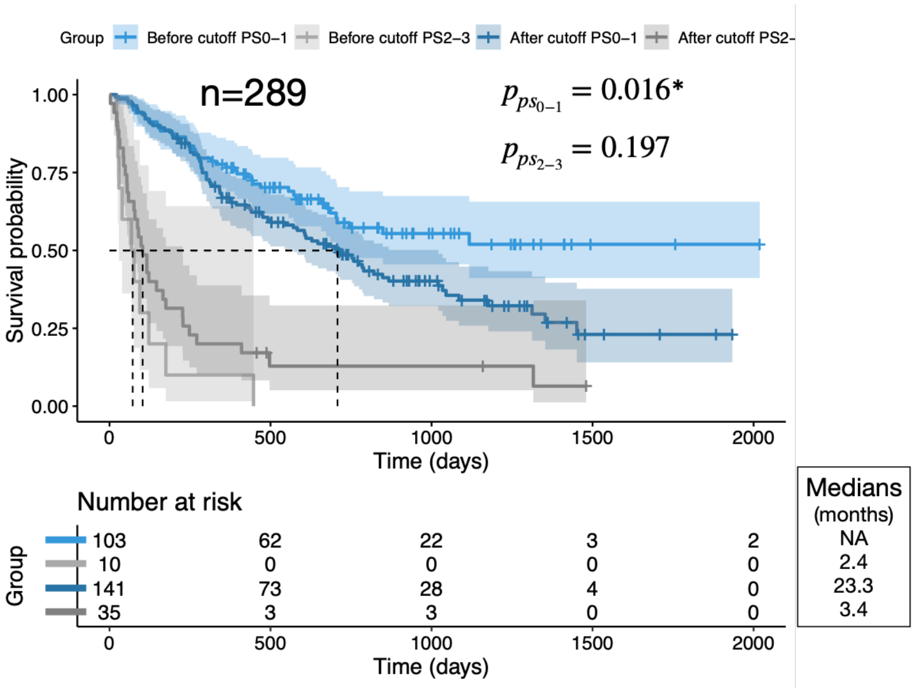


***Figure S4.*** Overall survival of the two infusion groups stratified by performance status, PS0-1 and 2-3, for the lung cancer patient subpopulation.

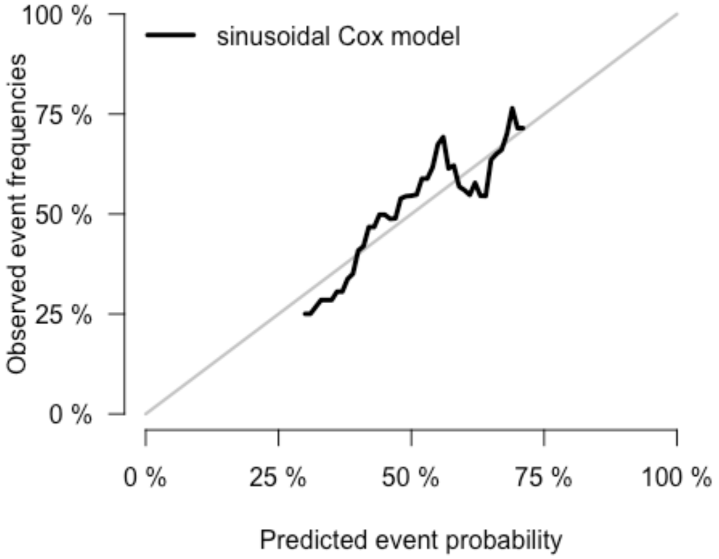


***Figure S5:*** Calibration plot assessing the sinusoidal Cox model's goodness-of-fit.

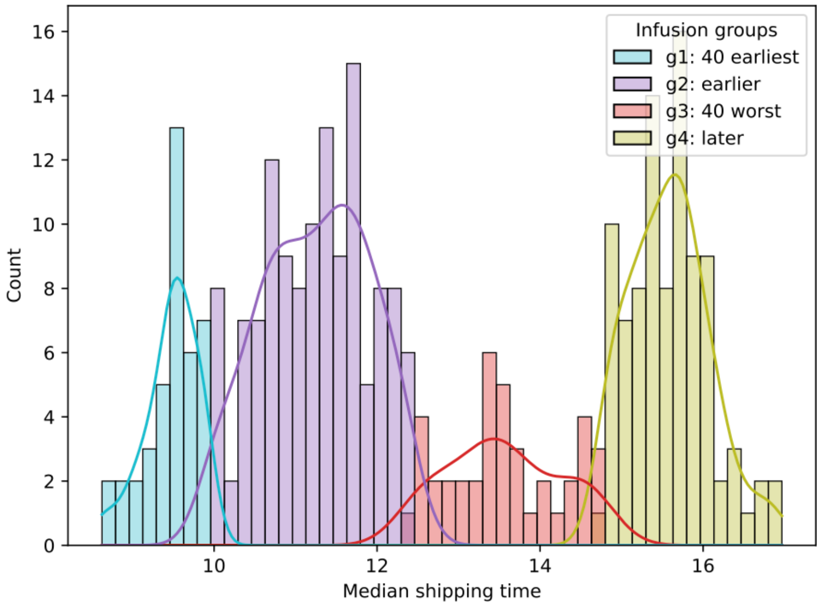


***Figure S6:*** Distribution and range of the shipping times by group.

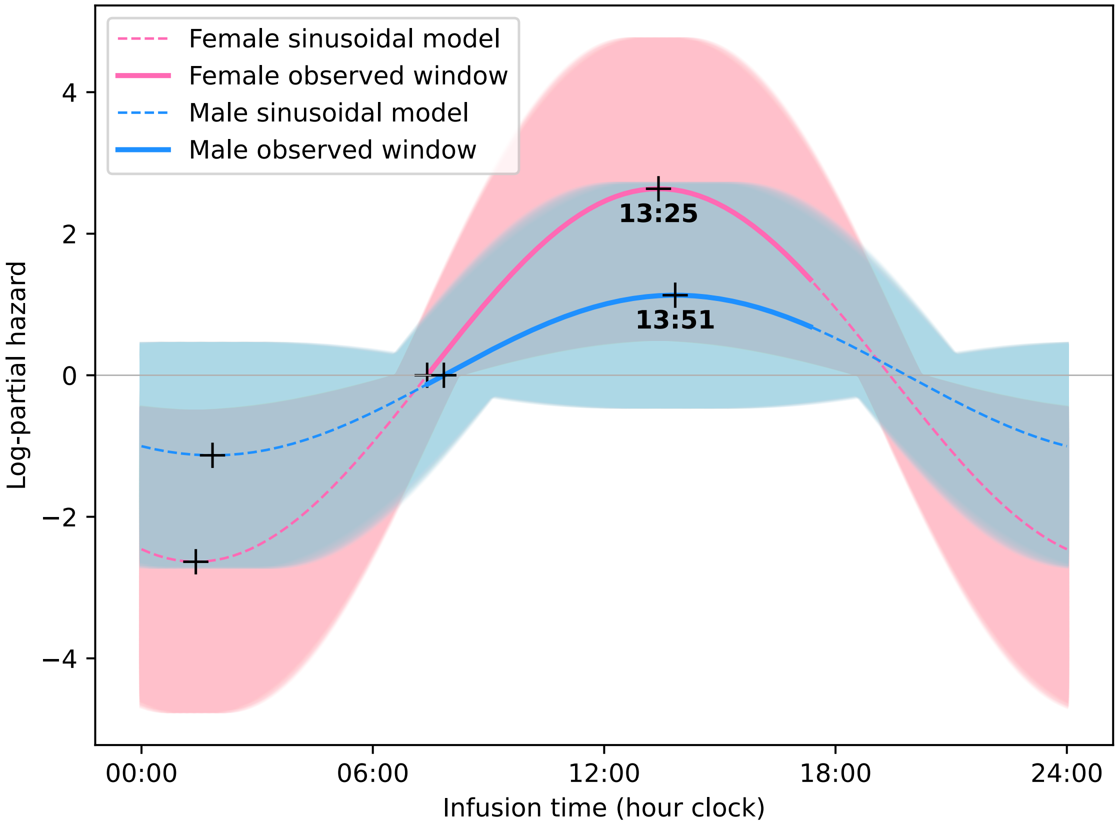


***Figure S7:*** Infusion timing model following a sinusoidal Cox regression by sex. Estimated amplitude A=2.64 and 1.13, and phase $\varphi$=13:25 and 13:51 time clock, for female and male patients, respectively. The shaded area represents the 95% CI for A (0.52–4.75) and (-0.45–2.78) and for $\varphi$ (12:37–14:12) and (12:09–15:33), for female and male patients, respectively. Observation window from 07:25 to 17:21 for both female and male patients.
