## Supplementary Information for "Early Morning Checkpoint Inhibitor Infusion and Overall Survival of Patients with Metastatic Cancer: an In-depth Chronotherapeutic Study"

### Sinusoidal Cox model

The sinusoidal Cox model for the infusion timing $\tau$ is given by

$$h(t) = h_0(t)\, e^{M+A\,cos(\tau\prime-\varphi\prime)} = h'_0(t)\, e^{A\,cos(\tau\prime-\varphi\prime)}$$

with $h'_0(t) = h_0(t)\, e^M$, $\tau' = \frac{2\pi}{T}\tau$ and $\varphi' = \frac{2\pi}{T}\varphi$, and denoting $A$ the amplitude, M the mesor, and $\varphi$ the phase.

Using the trigonometric formula $cos(a-b) = cos(a)\,cos(b) + sin(a)\,sin(b)$, the exponential term can be developed as such:

$$A\,cos(\tau' - \varphi') = b_1\,cos(\tau') + b_2\,sin(\tau')$$

with $b_1 = A\,cos(\varphi')$ and $b_2 = A\,sin(\varphi')$.
Given the Cox regression estimates $b_1$ and $b_2$, by definition, the ratio $b_2/b_1 = tan(\varphi')$, which allows to calculate the phase (in radians)

$$\varphi' = arctan(\frac{b_2}{b_1})$$

and the amplitude

$$A = \sqrt{b_1^2 + b_2^2}\ .$$

To obtain the phase in fraction of hours (and then in clock time), one can use the relation: $\varphi = \frac{T}{2\pi}\varphi'$.

### Hazard Ratio between worst and average timing

Using the sinusoidal Cox model, the hazard ratio between “worst” and “average” infusion time is calculated as the ratio between the model evaluated at $\tau' = \varphi'$ and at $\tau' = \varphi' - \frac{\pi}{2}$, where the cosine equals 1 and 0, respectively:

$$HR = \frac{h_0(t)e^{A\,cos(0)}}{h_0(t)e^{A\,cos(-\pi/2)}} = e^A.$$

The 95% CI is thus calculated by applying the exponential function to the lower and the upper bounds of the estimate $A$.
