## Supplementary Table S1 for "Early Morning Checkpoint Inhibitor Infusion and Overall Survival of Patients with Metastatic Cancer: an In-depth Chronotherapeutic Study"

| | | Missing | Overall | Morning | Afternoon | P-Value |
|---|---|---|---|---|---|---|
| n | | | 289 | 113 | 176 | |
| Median infusion time, median [Q1,Q3] | | 0 | 12.3 [10.8,15.3] | 10.6 [9.8,11.1] | 15.0 [12.9,15.8] | <0.001 |
| Age (years), mean (SD) | | 0 | 62.5 (10.3) | 61.5 (11.0) | 63.0 (9.9) | 0.233 |
| BMI, mean (SD) | | 2 | 23.9 (4.5) | 24.2 (4.8) | 23.8 (4.4) | 0.497 |
| Sex, n (%) | Female | 0 | 116 (40.1) | 43 (38.1) | 73 (41.5) | 0.648 |
| | Male | | 173 (59.9) | 70 (61.9) | 103 (58.5) | |
| Performance status, n (%) | 0-1 | 1 | 243 (84.4) | 102 (91.1) | 141 (80.1) | 0.020 |
| | 2-3 | | 45 (15.6) | 10 (8.9) | 35 (19.9) | |
| Immunoallergic history, n (%) | No | 1 | 269 (93.4) | 101 (90.2) | 168 (95.5) | 0.130 |
| | Yes | | 19 (6.6) | 11 (9.8) | 8 (4.5) | |
| Tumor PDL1 expression, n (%) | >1 | 168 | 63 (52.1) | 26 (54.2) | 37 (50.7) | 0.674 |
| | <1 | | 50 (41.3) | 20 (41.7) | 30 (41.1) | |
| | Not assessed | | 8 (6.6) | 2 (4.2) | 6 (8.2) | |
| Previous systemic treatment, n (%) | No | 1 | 107 (37.2) | 48 (42.9) | 59 (33.5) | 0.141 |
| | Yes | | 181 (62.8) | 64 (57.1) | 117 (66.5) | |
| Type of first immunotherapy, n (%) | Atezolizumab | 0 | 19 (6.6) | 9 (8.0) | 10 (5.7) | 0.331 |
| | Durvalumab | | 21 (7.3) | 11 (9.7) | 10 (5.7) | |
| | Nivolumab | | 82 (28.4) | 27 (23.9) | 55 (31.2) | |
| | Pembrolizumab | | 167 (57.8) | 66 (58.4) | 101 (57.4) | |
| Line of immunotherapy treatment, n (%) | 1st line | 1 | 108 (37.5) | 48 (42.9) | 60 (34.1) | 0.481 |
| | 2nd line | | 143 (49.7) | 49 (43.8) | 94 (53.4) | |
| | 3rd line | | 27 (9.4) | 11 (9.8) | 16 (9.1) | |
| | 4th line | | 9 (3.1) | 4 (3.6) | 5 (2.8) | |
| | 5th line | | 1 (0.3) | | 1 (0.6) | |
| Treatment with anti-inflammatory drugs, n (%) | No | 1 | 273 (94.8) | 102 (91.1) | 171 (97.2) | 0.046 |
| | Yes | | 15 (5.2) | 10 (8.9) | 5 (2.8) | |
| Next treatment, n (%) | No | 1 | 122 (42.4) | 49 (43.8) | 73 (41.5) | 0.796 |
| | Yes | | 166 (57.6) | 63 (56.2) | 103 (58.5) | |
| Type of next treatment, n (%) | Chemiotherapy | 123 | 150 (90.4) | 56 (88.9) | 94 (91.3) | 0.491 |
| | Surgery | | 1 (0.6) | 1 (1.6) | | |
| | Radiotherapy | | 14 (8.4) | 6 (9.5) | 8 (7.8) | |
| | Targeted therapy | | 1 (0.6) | | 1 (1.0) | |
