## Supplementary Table S2 for "Early Morning Checkpoint Inhibitor Infusion and Overall Survival of Patients with Metastatic Cancer: an In-depth Chronotherapeutic Study"

| | | Missing | Overall | Morning | Afternoon | P-Value all | P-Value PS0-1 | P-Value PS2-3 |
|---|---|---|---|---|---|---|---|---|
| n | | | 289 | 113 | 176 | | | |
| Response, n (%) | Progression | 5 | 84 (29.6) | 23 (21.1) | 61 (34.9) | 0.007 | 0.027 | 0.444 |
| | Stability | | 56 (19.7) | 18 (16.5) | 38 (21.7) | | | |
| | Partial response | | 107 (37.7) | 53 (48.6) | 54 (30.9) | | | |
| | Complete response | | 37 (13.0) | 15 (13.8) | 22 (12.6) | | | |
| Number of toxicities, n (%) | None | 0 | 172 (59.5) | 56 (49.6) | 116 (65.9) | 0.013 | 0.028 | 0.281 |
| | 1 | | 81 (28.0) | 42 (37.2) | 39 (22.2) | | | |
| | 2 | | 29 (10.0) | 10 (8.8) | 19 (10.8) | | | |
| | 3 | | 7 (2.4) | 5 (4.4) | 2 (1.1) | | | |
| Highest toxicity grade, n (%) | Grade 0 | 0 | 172 (59.5) | 56 (49.6) | 116 (65.9) | 0.015 | 0.033 | 0.281 |
| | Grade 1 | | 59 (20.4) | 30 (26.5) | 29 (16.5) | | | |
| | Grade 2 | | 39 (13.5) | 20 (17.7) | 19 (10.8) | | | |
| | Grade 3 | | 17 (5.9) | 7 (6.2) | 10 (5.7) | | | |
| | Grade 4 | | 2 (0.7) | | 2 (1.1) | | | |
| Number of infusions, median [Q1,Q3] | | 0 | 9.0 [4.0,21.0] | 11.0 [5.0,23.0] | 7.0 [4.0,20.0] | 0.016 | | |
| Treatment duration, median [Q1,Q3] | | 63 | 105.0 [42.0,278.0] | 127.5 [50.8,328.8] | 85.0 [42.0,210.2] | 0.035 | | |
| Discontinue Reason, n (%) | On going | 15 | 33 (12.0) | 15 (14.3) | 18 (10.7) | 0.182 | 0.307 | 0.632 |
| | Change/End treatment | | 28 (10.2) | 14 (13.3) | 14 (8.3) | | | |
| | Intolerance/Toxicity | | 20 (7.3) | 10 (9.5) | 10 (5.9) | | | |
| | Progression/Death/Palliative care | | 193 (70.4) | 66 (62.9) | 127 (75.1) | | | |
