## Supplementary Table S3 for "Early Morning Checkpoint Inhibitor Infusion and Overall Survival of Patients with Metastatic Cancer: an In-depth Chronotherapeutic Study"

| | | Missing | Overall | Morning | Afternoon | P-Value |
|---|---|---|---|---|---|---|
| n | | | 139 | 48 | 91 | |
| gender, n (%) | Female | 0 | 139 (100.0) | 48 (100.0) | 91 (100.0) | 1.000 |
| Performance status, n (%) | 0-1 | 0 | 115 (82.7) | 46 (95.8) | 69 (75.8) | 0.006 |
| | 2-3 | | 24 (17.3) | 2 (4.2) | 22 (24.2) | |
| Line of immunotherapy treatment, n (%) | 1st line | 0 | 52 (37.4) | 22 (45.8) | 30 (33.0) | 0.335 |
| | 2nd line | | 71 (51.1) | 23 (47.9) | 48 (52.7) | |
| | 3rd line | | 12 (8.6) | 2 (4.2) | 10 (11.0) | |
| | 4th line | | 4 (2.9) | 1 (2.1) | 3 (3.3) | |
| Number of infusions, median [Q1,Q3] | | 0 | 7.0 [4.0,21.0] | 11.5 [6.0,25.0] | 6.0 [3.0,19.5] | 0.006 |
| Treatment duration, median [Q1,Q3] | | 28 | 98.0 [42.0,290.5] | 129.0 [77.5,350.0] | 73.5 [40.8,217.5] | 0.028 |
| Discontinue Reason, n (%) | On going | 6 | 16 (12.0) | 8 (17.4) | 8 (9.2) | 0.254 |
| | Change/End treatment | | 14 (10.5) | 7 (15.2) | 7 (8.0) | |
| | Intolerance/Toxicity | | 12 (9.0) | 4 (8.7) | 8 (9.2) | |
| | Progression/Death/Palliative care | | 88 (66.2) | 27 (58.7) | 61 (70.1) | |
| | Other | | 3 (2.3) | | 3 (3.4) | |
| Response, n (%) | Progression | 2 | 44 (32.1) | 8 (17.4) | 36 (39.6) | 0.032 |
| | Stability | | 29 (21.2) | 9 (19.6) | 20 (22.0) | |
| | Partial response | | 43 (31.4) | 19 (41.3) | 24 (26.4) | |
| | Complete response | | 21 (15.3) | 10 (21.7) | 11 (12.1) | |
| Number of toxicities, n (%) | None | 0 | 75 (54.0) | 16 (33.3) | 59 (64.8) | <0.001 |
| | 1 | | 47 (33.8) | 27 (56.2) | 20 (22.0) | |
| | 2 | | 13 (9.4) | 2 (4.2) | 11 (12.1) | |
| | 3 | | 4 (2.9) | 3 (6.2) | 1 (1.1) | |
| Highest toxicity grade, n (%) | Grade 0 | 0 | 75 (54.0) | 16 (33.3) | 59 (64.8) | 0.003 |
| | Grade 1 | | 37 (26.6) | 20 (41.7) | 17 (18.7) | |
| | Grade 2 | | 16 (11.5) | 9 (18.8) | 7 (7.7) | |
| | Grade 3 | | 9 (6.5) | 3 (6.2) | 6 (6.6) | |
| | Grade 4 | | 2 (1.4) | | 2 (2.2) | |

| | | Missing | Overall | Morning | Afternoon | P-Value |
|---|---|---|---|---|---|---|
| n | | | 222 | 88 | 134 | |
| gender, n (%) | Male | 0 | 222 (100.0) | 88 (100.0) | 134 (100.0) | 1.000 |
| Performance status, n (%) | 0-1 | 0 | 184 (82.9) | 79 (89.8) | 105 (78.4) | 0.043 |
| | 2-3 | | 38 (17.1) | 9 (10.2) | 29 (21.6) | |
| Line of immunotherapy treatment, n (%) | 1st line | 1 | 75 (33.9) | 29 (33.3) | 46 (34.3) | 0.291 |
| | 2nd line | | 117 (52.9) | 42 (48.3) | 75 (56.0) | |
| | 3rd line | | 21 (9.5) | 12 (13.8) | 9 (6.7) | |
| | 4th line | | 7 (3.2) | 4 (4.6) | 3 (2.2) | |
| | 5th line | | 1 (0.5) | | 1 (0.7) | |
| Number of infusions, median [Q1,Q3] | | 0 | 8.0 [4.0,20.0] | 9.5 [4.8,20.5] | 7.0 [4.0,18.8] | 0.099 |
| Treatment duration, median [Q1,Q3] | | 45 | 98.0 [42.0,224.0] | 119.0 [45.5,321.5] | 75.5 [42.0,173.0] | 0.045 |
| Discontinue Reason, n (%) | On going | 20 | 17 (8.4) | 7 (9.0) | 10 (8.1) | 0.079 |
| | Change/End treatment | | 17 (8.4) | 9 (11.5) | 8 (6.5) | |
| | Intolerance/Toxicity | | 17 (8.4) | 11 (14.1) | 6 (4.8) | |
| | Progression/Death/Palliative care | | 150 (74.3) | 51 (65.4) | 99 (79.8) | |
| | Other | | 1 (0.5) | | 1 (0.8) | |
| Response, n (%) | Progression | 4 | 70 (32.1) | 22 (25.6) | 48 (36.4) | 0.065 |
| | Stability | | 45 (20.6) | 17 (19.8) | 28 (21.2) | |
| | Partial response | | 84 (38.5) | 42 (48.8) | 42 (31.8) | |
| | Complete response | | 19 (8.7) | 5 (5.8) | 14 (10.6) | |
| Number of toxicities, n (%) | None | 0 | 143 (64.4) | 53 (60.2) | 90 (67.2) | 0.404 |
| | 1 | | 51 (23.0) | 21 (23.9) | 30 (22.4) | |
| | 2 | | 24 (10.8) | 11 (12.5) | 13 (9.7) | |
| | 3 | | 4 (1.8) | 3 (3.4) | 1 (0.7) | |
| Highest toxicity grade, n (%) | Grade 0 | 0 | 143 (64.4) | 53 (60.2) | 90 (67.2) | 0.622 |
| | Grade 1 | | 32 (14.4) | 13 (14.8) | 19 (14.2) | |
| | Grade 2 | | 35 (15.8) | 16 (18.2) | 19 (14.2) | |
| | Grade 3 | | 11 (5.0) | 6 (6.8) | 5 (3.7) | |
| | Grade 4 | | 1 (0.5) | | 1 (0.7) | |
